# WELL-ED: Wellbeing and Education linkages in school-aged children - A protocol for a population-based register study and survey of adolescents

**DOI:** 10.64898/2026.06.06.26355053

**Authors:** Silja Kosola, Sanna Salonen, Jenni Miettinen, Iiris Hörhammer, Anna-Riikka Impiö, Satu M. Kumpulainen, Johanna Sergejeff, Saija Numari, Pirjo Laitinen-Parkkonen, Maria Tapola-Haapala, Elina Aaltio, Lena M. Thorn

## Abstract

**Introduction:** Education is a core social determinant of health for children and adolescents. Unfortunately, academic achievement, health, and wellbeing of adolescents have decreased in many developed countries in the past decade. The purpose of the Wellbeing and Education linkages in school-aged children (WELL-ED) study is to examine associations of school absences and academic achievement with use of school-based and community-based health and social welfare services. In addition, we will assess user experiences and multi-sector services pathways of school-aged children for a better understanding of how the service system could respond to the needs of children.

**Methods and analysis:** WELL-ED is a large population-based study that combines register data on school absences and educational support from municipalities with register data on healthcare and social service use collected from wellbeing services counties in Finland. The study cohort includes all children who attended mandatory education in public schools in Southern Finland in school year 2023-2024. A smaller cohort of adolescents in school year 8 was invited to complete a user experience survey. The primary outcomes of this study are related to equity of service use.

**Ethics and dissemination:** The Regional Committee on Medical Research Ethics of the Helsinki and Uusimaa Hospital District (2803/2024) has approved the WELL-ED study protocol. For the survey, adolescents in year 8 and parents of adolescents younger than 15 provided informed consent. Results will be published in peer-reviewed journals, summaries will be sent to participating municipalities and wellbeing services counties and press releases will be written on key findings.

## Introduction

Education, health, and wellbeing are closely intertwined. Education is strongly associated with health behaviours and life expectancy, and it may be considered one of the most important social determinants of health (1). To ensure students have the best possible opportunities for learning, more than 100 countries provide some form of school health services (2). The World Health Organization (WHO) and the United Nations Educational, Scientific and Cultural Organization (Unesco) have recognized the close connections of health and education and published several guideline documents to support the concept of health promoting schools (3).

Although many European countries provide free or subsidised education and health and welfare services to their children, recent years have seen increasing differences in academic achievement, health, and wellbeing among children and adolescents (4-6). The socio-economic status (SES) of the family bears more weight in education than before even in the Nordic countries which have previously been beacons of equity (7). In schools, especially absences due to illness are an increasing problem and teachers have very limited opportunities to intervene in them (8-10). Although school absences are associated with a lower educational level and poorer health in adulthood, systematic records on school absences and interventions to overcome them are still often lacking (11).

Increasing polarisation is also evident in health behaviour among adolescents: although binge drinking and smoking have decreased on a population level, socio-economic disparities in risky behaviour have increased (5, 12, 13). Furthermore, nearly all developed countries have witnessed a rise in mental health symptoms reported by adolescents. This rise began already prior to the COVID-19 pandemic, was exacerbated by social distancing measures, and has shown no signs of decline (14). Tackling polarisation and responding to adolescent mental health problems call for more targeted approaches, but concurrent, universal health promotion measures are also needed. In Finland, a social and healthcare service reform of historical proportions was undertaken in 2023 and gathered all social and healthcare services from more than 300 municipalities to 21 wellbeing services counties, while the city of Helsinki and Åland Islands are exceptions and organize their own services (15). Most true reforms, such as service redesign and integration, are still forthcoming. For example, a recent study supported the hypothesis that the concerns of teachers, parents and school nurses regarding primary school students are associated with the need and benefit of a consultation with a school physician (16), but no mechanism is in place to notify school healthcare providers of school absences or other concerns to ensure informational continuity of care (17).

In a critical assessment of school healthcare services in Finland, the evidence-base was modest. Some studies have estimated the benefits and potential harms of routine health checks conducted by school physicians in school years 1 and 5 (18), but no studies have targeted routine health checks in year 8. Furthermore, little is known about the health needs and access to services of children in other year levels at school. Meanwhile in child protection services, the proportion of adolescents in care began to increase before the pandemic and has shown no decline (19, 20). In many instances, child protection services have become the first point of contact although this position should belong to preventive and supportive family services. To increase effectiveness and to reduce inequities, policies and healthcare and welfare services should address structural and interpersonal discrimination that may further exacerbate socio-economic inequity (21). To date, no regularly measured quality indicators exist for service pathways of children and adolescents.

Researchers across disciplines have called for collaboration in research and development between professions and organisations because preventing social exclusion or finding solutions to help students with excessive school absences is impossible without cooperation of education, healthcare and social work professionals (8, 22). In an era of aging populations and decreasing birth rates, research combining these fields is of paramount importance.

### Study aims and hypotheses

The primary aim of this population-based study is, for the first time globally, to combine data from school administrative records with data on healthcare and social care service use to provide a holistic view of service use patterns and pathways for school-aged children. We also aim to estimate the needs, benefits and potential harms of routine health checks from the perspective of adolescents in school year 8.

More specifically, we aim to:

1. Measure the prevalence of school absences and sociodemographic and educational factors associated with them
2. Assess how well school healthcare services target students with problematic school absences (more than 10% of school year) or students in need of pedagogical support
3. Evaluate equity in healthcare and social service use based on the socio-economic area of the school
4. Map the multi-sector service pathways that start in school healthcare and wellbeing services, identify pathway discontinuities and propose quality indicators for them
5. Report experiences of care of students in school year 8 who have attended routine health checks with a school physician.

Our primary hypotheses are that 1) the prevalence of excessive school absences increases along with student age and that SES is associated with absences; and that 2) school healthcare service use has low correlation with school absences and demographic variables. Furthermore, we suspect that the care pathways of school children across student wellbeing services and social care services show room for improvement.

## Methods and analyses

### Study design and setting

The Wellbeing and Education linkages in school-aged children (WELL-ED) Study was designed together with representatives from the eight wellbeing services counties in Southern Finland (City of Helsinki and Wellbeing Services Counties of Central Uusimaa, East Uusimaa, Kymenlaakso, Päijät-Häme, South Carelia, Vantaa and Kerava, and Western Uusimaa).

In Finland, compulsory education begins the year a child turns seven years and more than 95% of children attend public schools. The first nine years of education follow a fairly uniform national core curriculum of basic education (23), with some locally weighted content, whereafter students enter secondary education where they choose either academic high school or vocational education. Assessment of academic achievement using numeric grades usually begins in the fourth year of basic education. In Finland, grades for school subjects range between 4 (fail) to 10 (excellent), with 8 (good) defined as generally reaching the set target for the school year (24). All municipalities use electronic school administration systems to track school absences, but no national register or system for follow-up of trends exists. Furthermore, no routine is in place for sharing school absence data with school healthcare providers.

During the nine-year basic education, wellbeing services counties are legally required to organize student health and wellbeing services (25-27). These services include mandatory annual health checks by a school nurse and three health checks by a school physician in school years 1, 5, and 8 (at ages 7-8, 11-12 and 14-15 years, respectively). The Finnish Supervisory Agency follows that the wellbeing services counties conduct the health checks as expected. Besides the health checks, school healthcare should also provide additional consultations based on the special needs of the students, but no systematic measure exists for this responsibility. In addition to school healthcare, the student health and wellbeing team must include school counsellor and school psychologist services. The primary aim of these services is to ensure a safe and inclusive school environment, but both professionals also meet individual students.

After ethics approval from the HUS Regional Committee on Medical Research Ethics (2803/2024), we applied for research permits from the eight wellbeing services counties and the corresponding 50 municipalities of Southern Finland. All wellbeing services counties and 38 (76%) municipalities granted permission to conduct the study. In all, register data of 188,016 children aged 7-15 (90% of the respective age group in the study area and 36% of the respective age group in Finland) are included in the study (please see Figure 1).

**Figure 1.**
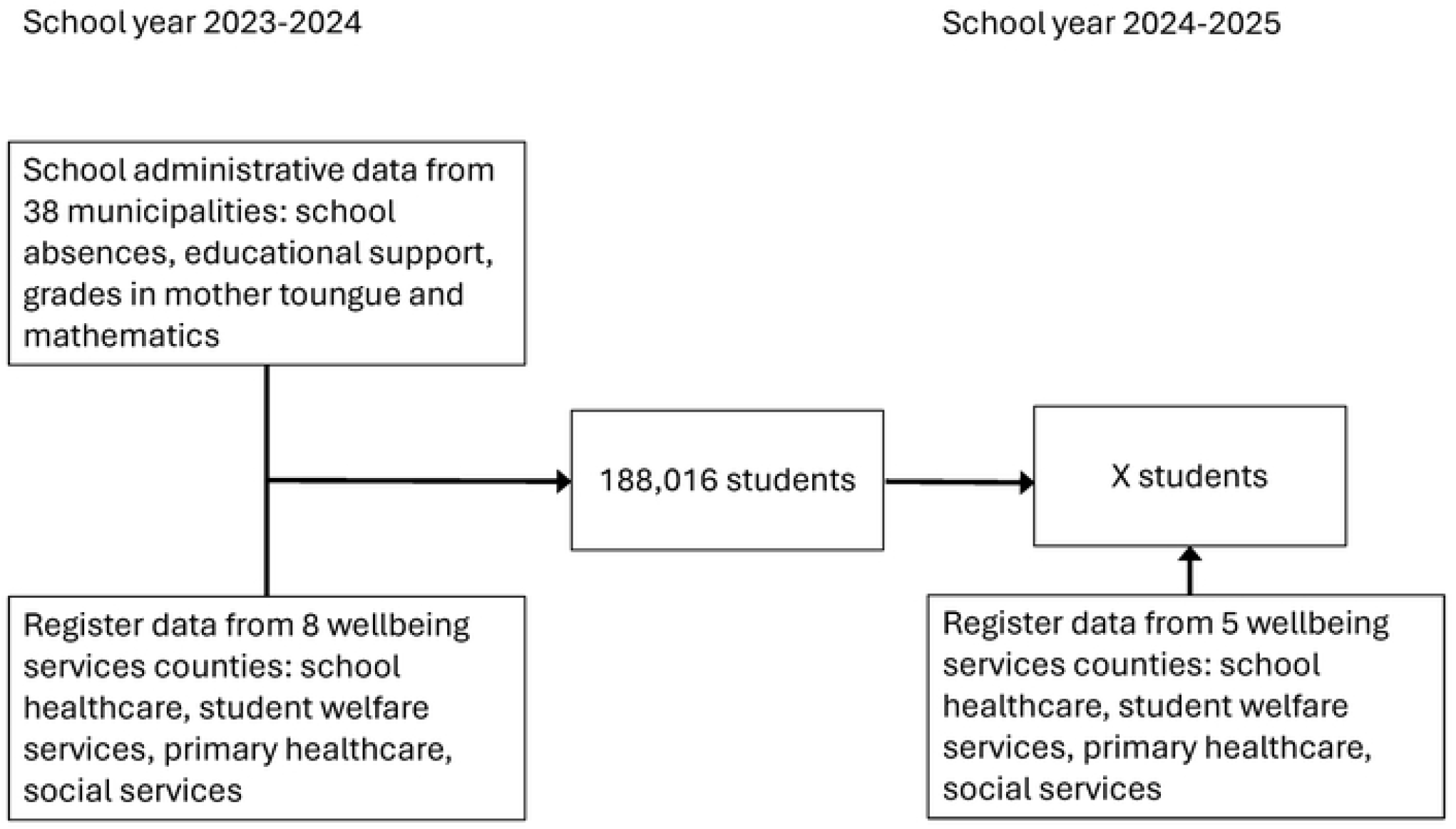
Flowchart of register data collection.

### Register data

Individual-level register data for the school year 2023-2024 were derived from the electronic school administration system Primus used by municipalities and the different electronic client and patient records of the wellbeing services counties (Table 1). Similar data from client and patient records will also be collected for the school year 2024-2025 to enable more in-depth analysis of service pathways and longitudinal analyses of service efficacy. This data collection is expected to be completed by August 2026.

**Table 1.**
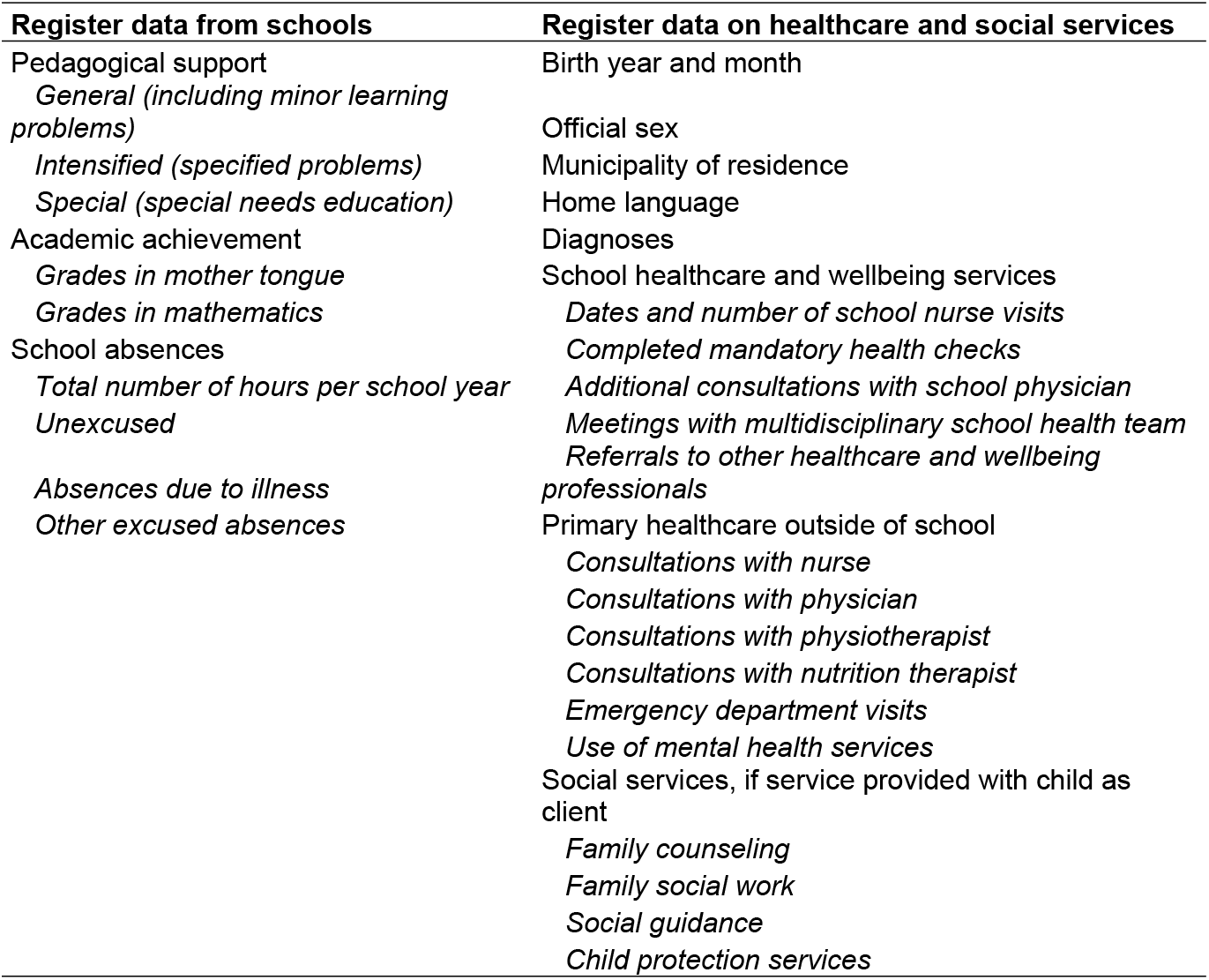
Data collected from municipalities (school data) and wellbeing services counties (healthcare and social services).

| Register data from schools | Register data on healthcare and social services |
| --- | --- |
| Pedagogical support | Birth year and month |
| *General (including minor learning problems)* | Official sex |
| *Intensified (specified problems)* | Municipality of residence |
| *Special (special needs education)* | Home language |
| Academic achievement | Diagnoses |
| *Grades in mother tongue* | School healthcare and wellbeing services |
| *Grades in mathematics* | *Dates and number of school nurse visits* |
| School absences | *Completed mandatory health checks* |
| *Total number of hours per school year* | *Additional consultations with school physician* |
| *Unexcused* | *Meetings with multidisciplinary school health team* |
|  | *Referrals to other healthcare and wellbeing professionals* |
| *Absences due to illness* | Primary healthcare outside of school |
| *Other excused absences* | *Consultations with nurse* |
|  | *Consultations with physician* |
|  | *Consultations with physiotherapist* |
|  | *Consultations with nutrition therapist* |
|  | *Emergency department visits* |
|  | *Use of mental health services* |
|  | Social services, if service provided with child as client |
|  | *Family counseling* |
|  | *Family social work* |
|  | *Social guidance* |
|  | *Child protection services* |

### Data on pedagogical support, school grades, and school absences

All public schools of all municipalities in Southern Finland use the same school administration system, Primus (Visma Software International AS), to record pedagogical support and school grades, and to track school absences. We collected data on educational support, school grades in mathematics and Finnish/Swedish (depending on which of the two national languages the students studied as their mother tongue or primary language at school), and school absences. In the school year 2023-2024, pedagogical support was based on a three-tiered system: 1) general support for all students, including students with minor learning problems, 2) intensified support for specified problems, and 3) special support or special needs education (28). Grades in mother tongue and mathematics were dichotomized: grade 7 or lower indicated that a student was not achieving the goals set for the respective school year. Teachers mark absences into the system using three different codes: absent with permission from parents/guardians (e.g. illness or family vacation), absent with permission from school (e.g. participation in tutoring), or absent without permission. In the last case, a notification is sent via the Primus digital user interface to the parents/guardians who have the possibility to mark the absence as excused due to illness or to leave it unexplained/unexcused. Some municipalities only record the total amount of absences without distinction between absence types, whereas others categorize absences as unexcused absences (also known as truancy), illness absences, and other excused absences. We collected both the total hours of absences for the school year, and when available, the hours of absences in the three categories (unexcused, illness absences, and other excused absences). To facilitate comparisons between different age groups, we will calculate two variables based on the national core curriculum (28): whether a student was absent for more than 10% of the school year, which is often considered a threshold for problematic school absences (29), and whether a student was absent for more than 20% of the school year, indicating a risk of educational dropout.

### Healthcare and social services data

At baseline (2023-2024), most of the participating wellbeing services counties of Southern Finland provided primary healthcare (but no hospital care) and social services. They used ten different client and patient record systems, and data retrieval had to be tailored to each one. The retrieved healthcare data included the following: national personal identification code (unique code provided to each individual after birth or if a legal resident has grounds for registration), birth year and month, official sex, municipality of residence, home language, all diagnoses, dates of visits to the school nurse, completed mandatory health checks, additional consultations with the school physician, possible meetings of a multidisciplinary health and wellbeing team, referrals to another health and wellbeing professional; and outside of schools in the public healthcare centres, consultations with a nurse, physician, physiotherapist, or nutrition therapist; emergency department visits, and treatment at mental health services. From social service records, we collected data on service use in family counselling, family social work, social guidance, and child protection services, if services were provided based on the child as a client.

### School socioeconomic area

In Finland, municipalities are primarily responsible for organizing compulsory education, and only 3-5% of Finnish students attend private schools during basic education (30). Because collection of household income data is inconceivable in this study, we will gather information on the socioeconomic area of the schools from Statistics Finland (40). A school-area index of socioeconomic disadvantage (SEI) combines three indicators at the postal code level: 1) the proportion of households belonging to the lowest income category, proportion of 25-64-year-old residents without secondary education, and the unemployment rate among residents aged 16-64. These indicators are summed and thereafter their mean is calculated. The sum index is proportional to the average of the municipality or wellbeing services county with the value 100 representing the average score. A value above 100 indicates that the SES of the postal code area is below the municipality average and a value below 100 implies that SES is above the municipality average. This index has previously been used by the Urban Research and Statistics Unit of the City of Helsinki.

### Data archive and management

After retrieval, all register data will be transferred to HUS Acamedic, an audited, secure processing environment for the study of sensitive social and healthcare data. The environment is accessible to registered research group members via a web browser. Due to the Act on the Secondary Use of Personal Data, eight separate data processing environments were created for the WELL-ED dataset, one for each health and wellbeing services county.

A data manager combines data from Primus and the different client and patient records using the unique personal identifier. After completing data combinations, all datasets are pseudonymized using a separate key. Researchers only have access to pseudonymous datasets.

### Survey data

#### Eligibility criteria

Inclusion criteria for the adolescent survey were that they: were at least 15 years old (according to Finnish research legislation, 15-year-olds can consent to participate in survey studies without parental consent), or if they were younger, parents provided consent; had attended a routine health check conducted by a school physician or had been offered one; attended one of the participating schools; and spoke at least one of the languages (Finnish, Swedish, English, Estonian, Russian, Arabic or Somali; most common home languages in the study area) in which the study survey was available. Surveys were translated by professional translators.

#### Sample size and power calculations

To ensure sufficient sample size, we conducted two power calculations, one for binary and one for continuous measures. First, we wished to detect small-to-moderate differences in binary outcomes (absolute differences of approximately 8-10 percentage points, corresponding to odds ratios of roughly 1.4-1.6) with 80 % power (α = 0.05, two-sided). For continuous outcomes, we wished to detect small, standardized effect sizes (Cohen’s d = 0.20). For both assumptions to be true, we would need a sample size of 800 participants. Considering the common recruitment rate of 30% among the target age group, we aimed to contact 2700 adolescents.

#### Participant recruitment

After ethics approval, we contacted 37 schools of which 19 agreed to participate. Three members of our study group visited the schools between August 15 and October 10, 2025, and provided year 8 students with both written and oral information about the study. Information was also available as short videos in the study languages. Participation was completely voluntary, and students were allowed to respond to the study survey during school hours. Surveys were available via QR codes in the seven different languages and responses were collected via Research Electronic Data Capture (REDCap), a secure digital survey tool provided by the University of Helsinki, Finland.

#### Survey content

The Patient Enablement Instrument (PEI) is a validated six-item, patient-reported outcome tool that evaluates a patient’s ability to understand, manage and cope with their condition after a consultation (32). It has been validated as a quality measure in primary healthcare in several countries, including Finland and Sweden (33, 34). Total PEI scores thus range from -6 to 12 or 0 to 12, respectively. We found no previous studies using PEI in adolescent populations.

In addition to the PEI, the survey included several binary items (e.g. “Prior to the meeting with your school doctor, did you have any issues you wished to discuss?”) as well as space for open responses.

### Analysis plan

Due to data protection regulations, all statistical analyses will be conducted separately for each of the eight wellbeing services counties and then combined and compared in meta-analyses.

We will adhere to the Strengthening the Reporting of Observational Studies in Epidemiology (STROBE) when reporting quantitative findings. Demographic and descriptive categorical data will be reported as numbers and percentages, and continuous variables as means with standard deviations (SD), or in case of skewed distribution, medians with interquartile ranges (IQR). Pearson and Spearman correlations will be calculated for binomial associations as appropriate. Multivariable regression analyses and mixed-effects models will be conducted, and data will be clustered according to municipality and schools, when appropriate.

ORs will be calculated to estimate the predictive value of age, home language, need for pedagogical support, and absences on appointments with school physicians and other school health and wellbeing professionals. To study the volume, frequency, and sequence of service use in the health and wellbeing service system, Sankey diagrams will be used to explore the flow of at-risk children across different social and healthcare settings and to probe suggestions for multi-sector quality indicators from the sector experts. Regression analysis will be used to identify statistical differences in service trajectories between sub-groups. To propose system-level quality indicators sensitive to resource allocation, cost estimates will be assigned to service use. Standard national cost estimates will be used where available and complemented by local estimates as needed.

### Patient and public involvement

The WELL-ED study design was cocreated with staff from all participating wellbeing services counties who also named a member to the steering group.

The survey for year 8 students was developed together with the Youth Research Council (YRC) of the New Children’s Hospital, which is part of the Helsinki University Hospital, Finland. The YRC is composed of nine voluntary adolescents, aged 15-17 years, who have been trained to serve on the YRC. The only fixed element of the survey was the PEI, some other questions were presented to the YRC as rough drafts and sent to the YRC two weeks before their meeting. Two researchers participated in the meeting with the YRC. The researchers mostly listened to the YRC members discuss the comprehensibility and content of the survey but also responded to questions from the YRC. The YRC was satisfied with the final survey and especially highlighted their support for the study subject, experiences of care of young people.

## Data Availability

No datasets were generated or analysed during the current study. All relevant data from this study will be made available upon study completion.

## Ethics and dissemination

The WELL-ED study conforms to the Declaration of Helsinki. The Regional Committee on Medical Research Ethics of the Helsinki and Uusimaa Hospital District approved the study protocol (2803/2024). For use of the register data, research permits were acquired from each participating organization and municipality. For the survey, adolescents and their parents/guardians received both verbal and written information on the purpose of the study. In Finland, adolescents aged 15 or older may give consent to participate in survey studies without approval from their parents. No compensation was provided for study participants.

Study results will be presented at scientific meetings and published in international, peer-reviewed journals, with a preference for open-access publication. After publication, summaries of study results will be disseminated via the research project’s website, the steering group, and participating municipalities. News media will also be notified.

In the coming years, we aim to build a curated repository of the WELL-ED register data together with researchers from the University of Turku, Finland. This repository will be stored in the FIONA remote access system, a secure environment for processing research data provided by Statistics Finland.

## Acknowledgements

The authors wish to thank research coordinator Rhea Paajanen from the Western Uusimaa Wellbeing Services County, Marjo Alatalo from the City of Helsinki, Elina Eeva from the Wellbeing Services County of Vantaa and Kerava, Eija Kinnunen and Kirsi Kuusinen-James from the Wellbeing Services County of Päijät-Häme, Päivi Mattila from the Wellbeing Services County of Kymenlaakso, Sirkka Pennanen from the Wellbeing Services County of South Carelia, Anni Pasuri from the Wellbeing Services County of Eastern Uusimaa, data manager Petri Vänni, members of the Youth Research Council of the Helsinki University Hospital, representatives of the municipalities who granted research permits and participated in data retrieval, and Emmi Turunen who helped with the setup of the secure data archive.

